# Antibiotic usage and susceptibility patterns in Uncomplicated UTI in a Tertiary Hospital in South India

**DOI:** 10.1101/2023.07.02.23292129

**Authors:** Christy John, Nithyananda Chowta, Sathish Rao

## Abstract

**Purpose:** Urinary tract infections (UTI) are common infections in otherwise healthy young women and there is considerable heterogeneity in antibiotic prescribing practices contributing to increased health expenditure, hospitalization, and ‘collateral damage’ with the unprecedented use of fluoroquinolones and beta-lactams leading to the increase in MRSA strains and gram-negative bacilli. This study was designed to study the appropriateness of empirical antimicrobial therapy with clinical outcomes among outpatients and analyze antibiotic susceptibility patterns.

**Methods:** A longitudinal study involving women clinically diagnosed with uncomplicated UTI as per IDSA guidelines across a study period of 18 months was conducted from 2008-2020. Antibiotic appropriateness was analyzed with respect to IDSA guidelines with subgroup analysis of culture-proven UTI.

**Results:** Among 105 cases of uncomplicated UTI, fluoroquinolones were prescribed the most (41%) followed by Beta-Lactams (30.5%). Choice of antimicrobial agent was appropriate in 60 (57%) cases and duration in 30 (28.5%) cases. Subgroup Analysis of 25 culture-proven cases revealed most common organism isolated was E. coli (60%) with prevalence of MDR organisms being 36%. The susceptibility pattern revealed similar levels of resistance between fluoroquinolones (38%), beta-lactams (36%), Nitrofurantoin(32%), Trimethoprim-sulfamethoxazole (32%), and Fosfomycin (20%) with clinical cure rates non-inferior among those prescribed the latter group of antibiotics (84.2% vs 96.6%)

**Conclusion:** The spectrum of uropathogens in our clinical setting is evolving with a substantial rise in MDR pathogens due to inappropriate antibiotic prescribing practices. The use of antibiotics such as Nitrofurantoin, Trimethoprim-sulfamethoxazole, and Fosfomycin in accordance with local antibiograms must be encouraged as empirical therapy for uncomplicated UTI.

## Introduction

Urinary Tract Infections (UTI) are among the most common infections occurring in the general population around the world, especially among young women in the reproductive age group. It is also one among the most common indications for a short course of oral antibiotics in otherwise healthy persons. Recent estimates suggest a global health expenditure upwards of 6 Billion USD for the management of urinary tract infections.[1]

UTI has an annual incidence of approximately 12% in women, with half of all women reporting at least a single episode of urinary tract infection by 32 years of age.[2] Among healthy young women with cystitis, UTI has been found to recur even in 25% of young healthy women within 6 months after the first infection with risk of subsequent infection rising following multiple episodes of urinary tract infection.[3–6] The incidence of acute uncomplicated pyelonephritis is much lower than that of cystitis.[7]

Uncomplicated UTI generally comprises of Episodes of acute cystitis and pyelonephritis occurring in healthy premenopausal, non-pregnant women with no history suggestive of an abnormal urinary tract. Usually, all other urinary tract infections are classified as complicated.[8] This distinction has implemented in clinical diagnoses and to decide on outpatient or in-hospital based care. It is also used to guide the choice and duration of antibiotics in persons with urinary tract infection.

Uncomplicated UTI includes acute uncomplicated cystitis and acute uncomplicated pyelonephritis.

Uncomplicated cystitis is defined by typical lower urinary tract symptoms such as dysuria, frequency of sudden onset, in the absence of vaginal discharge.[7] Acute uncomplicated pyelonephritis is characterized by symptoms of fever with chills, flank pain and renal angle tenderness.

Persons with uncomplicated UTI are usually treated with a short course of empirical, oral antibiotic therapy. However, a significant change in patient outcomes have been observed with increasing hospitalization and recurrence of UTI. This has been attributed to the evolution of the spectrum of commonly isolated uropathogens with the emergence of multi-drug resistant strains.[8]

This phenomenon has been exacerbated by the inappropriate antibiotic therapy, and the dearth of local antibiotic susceptibility patterns to guide therapy. The paucity of guidelines by governing bodies regarding antibiotic usage in UTI has led to increased use of certain antibiotics such as Fluoroquinolones, Beta-Lactams.

This has contributed to increasing prevalence of microbial strains resistant to these broad-spectrum antibiotics, thereby decreasing their potential for other infections.

Recent guidelines by international bodies such as IDSA warn against such “collateral damage” and advocate the use of agents such as Nitrofurantoin (NFT), Trimethoprim-Sulfamethoxazole (TMP-SMX), Fosfomycin and Pivmecillinam (in available countries) in UTI. However, there is a dearth of local antibiograms to substantiate their use in the South Indian clinical setting.

The lack of standardization of therapeutic regimen in the Indian setting has contributed to burgeoning multi-drug resistant pathogens such as Methicillin Resistant Staphylococcus Aureus (MRSA) and Gram-Negative Bacilli such as Pseudomonas. Therefore, antibiotic stewardship is required to mitigate such trends.

## Declarations

### Ethics Approval

The study was approved by the Institutional Review Board at KMC Mangalore.

### Consent to Participate

All participants had provided their informed consent prior to being enrolled into the study.

### Consent for Publication

All participants had provided their informed consent for publication of the findings of the study.

### Availability of Data and Material

The data supporting the findings of the study are available from the corresponding author upon request.

### Code Availability

Not applicable

## Patients and Methods

### Study Setting

This was a longitudinal study conducted in the outpatient departments of Kasturba Medical College (KMC) teaching hospitals, Mangalore, Karnataka, India from 1^st^ September 2018 to 31^st^ March 2020. KMC affiliated hospitals comprise of KMC Attavar and KMC Ambedkar Circle hospitals accounting for 850 beds.

### Case Selection

All adult, female patients attending the outpatient department of the affiliated hospitals who were clinically diagnosed to have uncomplicated urinary tract infection were invited to participate in the study. The patients were considered for possible enrolment into the study by screening their clinical history. All cases with coexisting conditions contributing to an altered host response to infection such as Diabetes Mellitus, or with a recent history of antimicrobial use or urological interventions were excluded. All cases were started empirically on oral antimicrobial therapy, and testing for urine culture and susceptibility was done at the attending physician’s discretion and all cases were asked to follow-up after one week since their visit.

### Data Collection

Clinical and laboratory data of all cases were collected prospectively from interviews and medical records. We recorded data on age, symptoms and duration on presentation, empirical antimicrobial agent initiated with urine susceptibility results and clinical outcomes on follow-up after a week as depicted in Figure 1.

**Figure 1.**
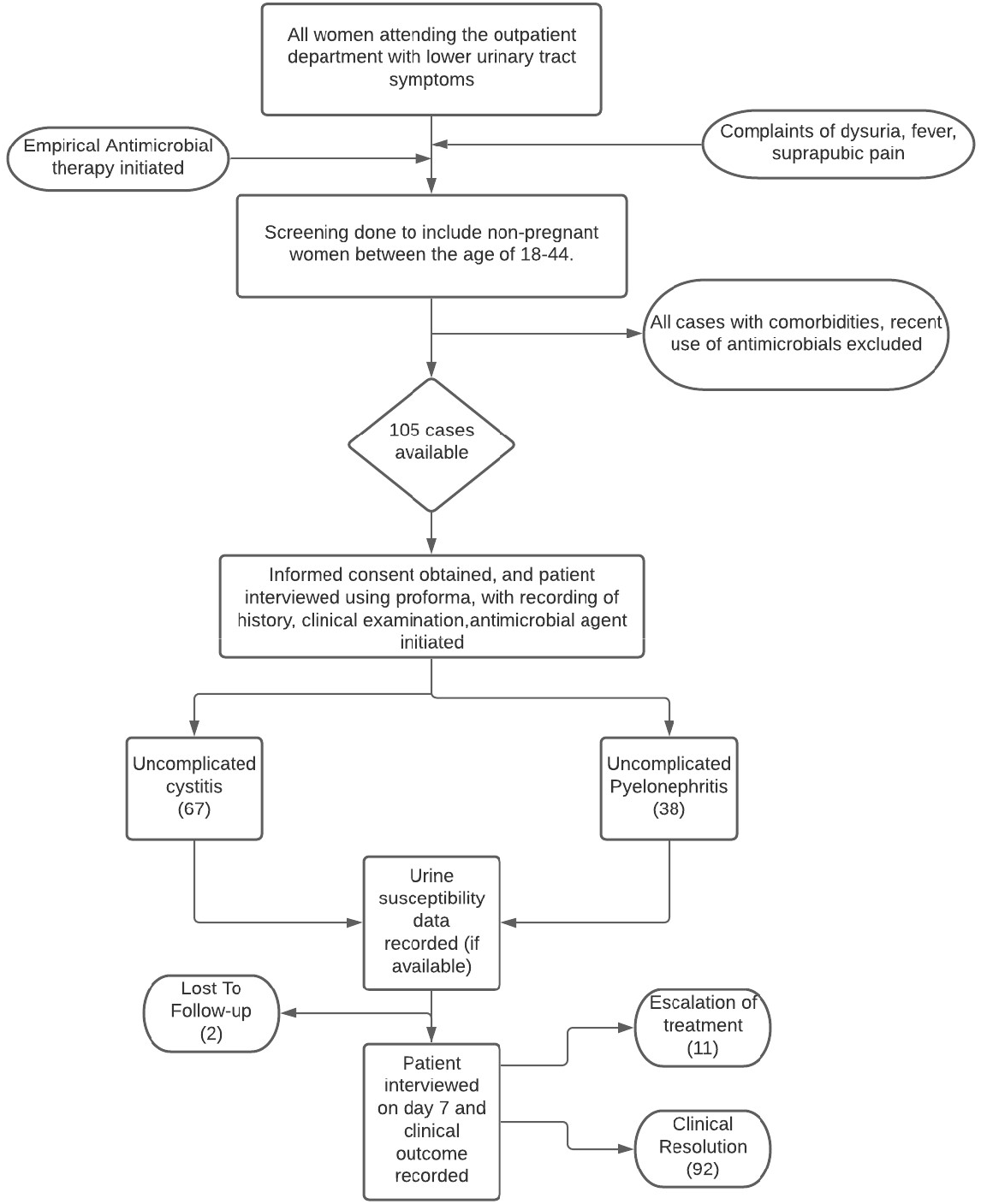
Methodology. Screening of all cases was done prior to enrollment and obtaining consent (n=105). All cases were assessed clinically by questionnaire method and clinically classified to have Uncomplicated Cystitis and Pyelonephritis. Susceptibility Data was collected, and clinical outcomes were determined on follow-up after 1-week

### Definitions

Uncomplicated UTI was defined as laid down by the most recent international guidelines as, “*Acute, sporadic or recurrent lower (uncomplicated cystitis) and/or upper (uncomplicated pyelonephritis) UTI, limited to non-pregnant women with no known relevant anatomical and functional abnormalities within the urinary tract or comorbidities*.”[8] Cases of cystitis were considered to have typical lower urinary symptoms such as dysuria, frequency. Cases having fever and flank pain were assumed to have pyelonephritis.

Clinical cure was attained if the patient had resolution of symptoms on follow-up after 1 week. If the patient had persistence of symptoms, they would be termed as requiring escalation of treatment. Urine cultures found to have a bacterial count of ≥ 10^3^ CFU/ml were deemed positive and evaluated further for identification of organism with antibiotic susceptibility, which were performed according to the local laboratory standards.

The antimicrobial agent prescribed were defined to be either a Class I or a Class II agent. Class I consisted of NFT, TMP-SMX and Fosfomycin whereas Class II consisted of Fluoroquinolones and Beta Lactam antimicrobials.

### Statistical Analysis

All cases were divided into two groups, Uncomplicated cystitis and Uncomplicated pyelonephritis, which were compared for presenting symptoms, appropriateness of empirical therapy, and clinical outcomes. The patients lost to follow up were excluded from outcome analysis.

Antibiotic susceptibility data was interpreted using descriptive statistics. Chi square test were used to compare continuous variables between the two groups with a p-value < 0.005 to denote statistical significance. Descriptive statistics were calculated using SPSS (Statistical Package for the Social Sciences) version 11.5.

## Results

A total of 105 women diagnosed to have uncomplicated UTI were enrolled into the study out of which 103 women were available for follow-up after 1 week. 67 women (63.8%) were diagnosed to have uncomplicated cystitis and 38 women (36.2%) to have uncomplicated pyelonephritis. The urine culture positivity noted in our study was 31.3%.

### Demography and clinical features

Majority of the women enrolled were in the age group of 21-30 years (67.6%) with a mean age of 26.3 ± 5.8 years. The most common symptom reported being dysuria (78.1%) followed by increased frequency of voiding urine (55.2%). Symptoms of fever and lower abdominal pain were noted more in cases diagnosed to uncomplicated pyelonephritis in a statistically significant manner. Table 1 compares the demographic features and clinical features on presentation between the cases of uncomplicated cystitis and pyelonephritis.

**Table 1.** Symptomatology. Patient characteristics in relation to classification have been described (n = 105); all patients who were included in the study and divided into cases of uncomplicated cystitis and pyelonephritis Parenthesis contains percentages a – described in mean ± SD

|  | **Total**<br>(n = 105) | **Uncomplicated**<br>**Cystitis**<br>(n = 67) | **Uncomplicated**<br>**Pyelonephritis**<br>(n = 38) | **Chi Square test/**<br>**Fisher's exact**<br>**test**<br>(p < 0.05) |
| --- | --- | --- | --- | --- |
| **Age** |  |  |  |  |
| All | 26.3 ± 5.8 | 26.8 ± 6.2 | 25.4 ± 5.0 |  |
| 20 and below | 11 | 8 (72.7) | 3 (27.3) | 0.166 |
| 21 - 30 | 71 | 41 (57.7) | 30 (42.3) |  |
| Above 30 years | 23 | 18 (78.3) | 5 (21.7) |  |
| **Dysuria** | 82 (78.1) | 55 (67.1) | 27 (32.9) | 0.189 |
| **Frequency**<br>**of micturition** | 68 (64.7) | 40 (69) | 18 (31) | 0.222 |
| **Lower**<br>**abdominal pain** | 40 (38) | 13 (32.5) | 27 (67.5) | <0.01 |
| **Fever** | 45 (42.8) | 14 (31.1) | 31 (68.9) | <0.01 |
| **Duration** |  |  |  |  |
| All | 3.5 ± 1.3 | 3.5 ± 1.3 | 3.6 ± 1.4 |  |
| 1 - 2 days | 11 | 16 (66.7) | 8 (33.3) | 0.939 |
| 3 - 4 days | 71 | 33 (63.5) | 19 (36.5) |  |
| 5 - 7 days | 23 | 18 (62.1) | 11 (37.9) |  |

### Microbiology

The most common organism isolated was E. coli (60%), with a significant proportion being ESBL producing (24%). The overall prevalence of MDR organisms was 36%. Figure 2 shows the uropathogens isolated.

**Figure 2.**
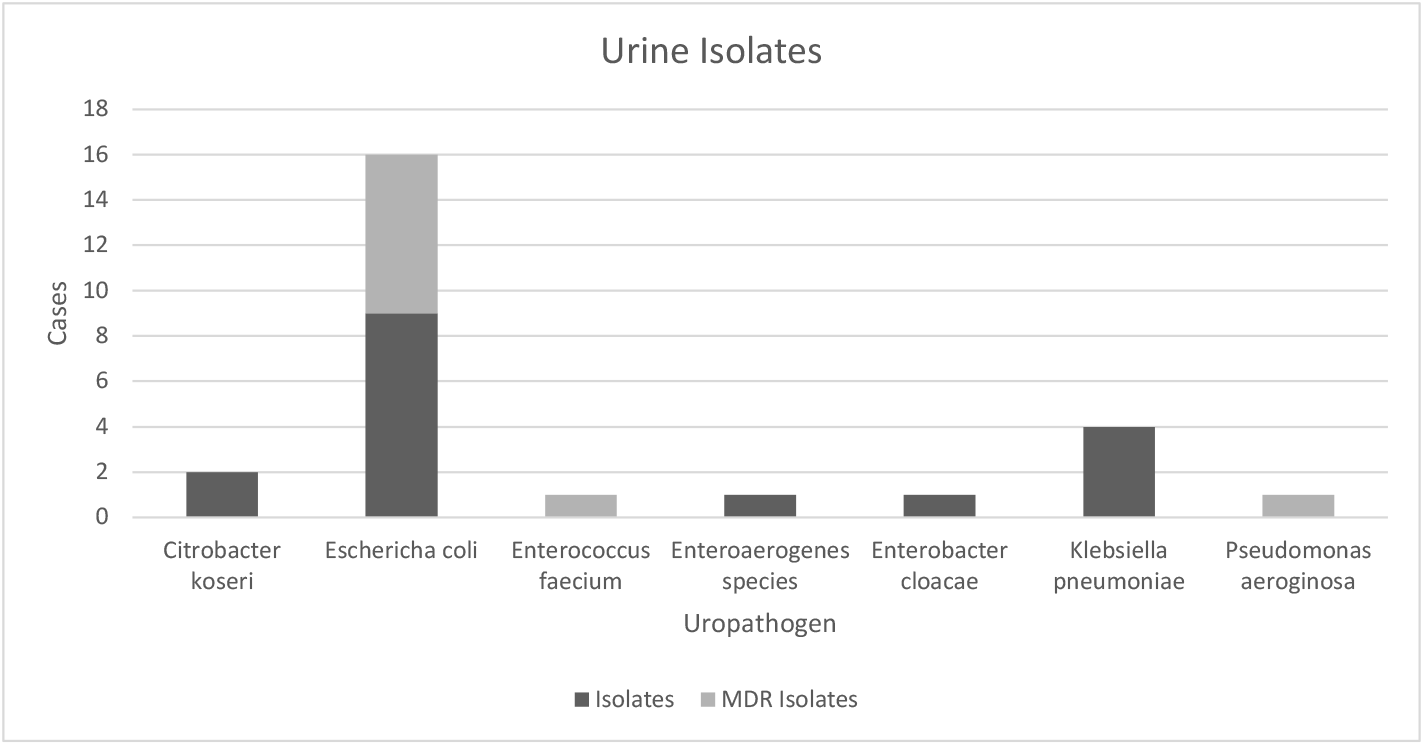
Urine isolates. Urine culture data revealing all isolated uropathogens with MDR strains MDR – Multi-Drug Resistant Organisms

### Appropriateness of Therapy

Many of the participants (71.4%) were prescribed a Class II antimicrobial agent; the most common one being Norfloxacin (23.8%). Overall, the most common group of antibiotics prescribed were Fluoroquinolones (41.1%) followed by Beta Lactams (30.5%). This is represented in Table 2.

**Table 2.**
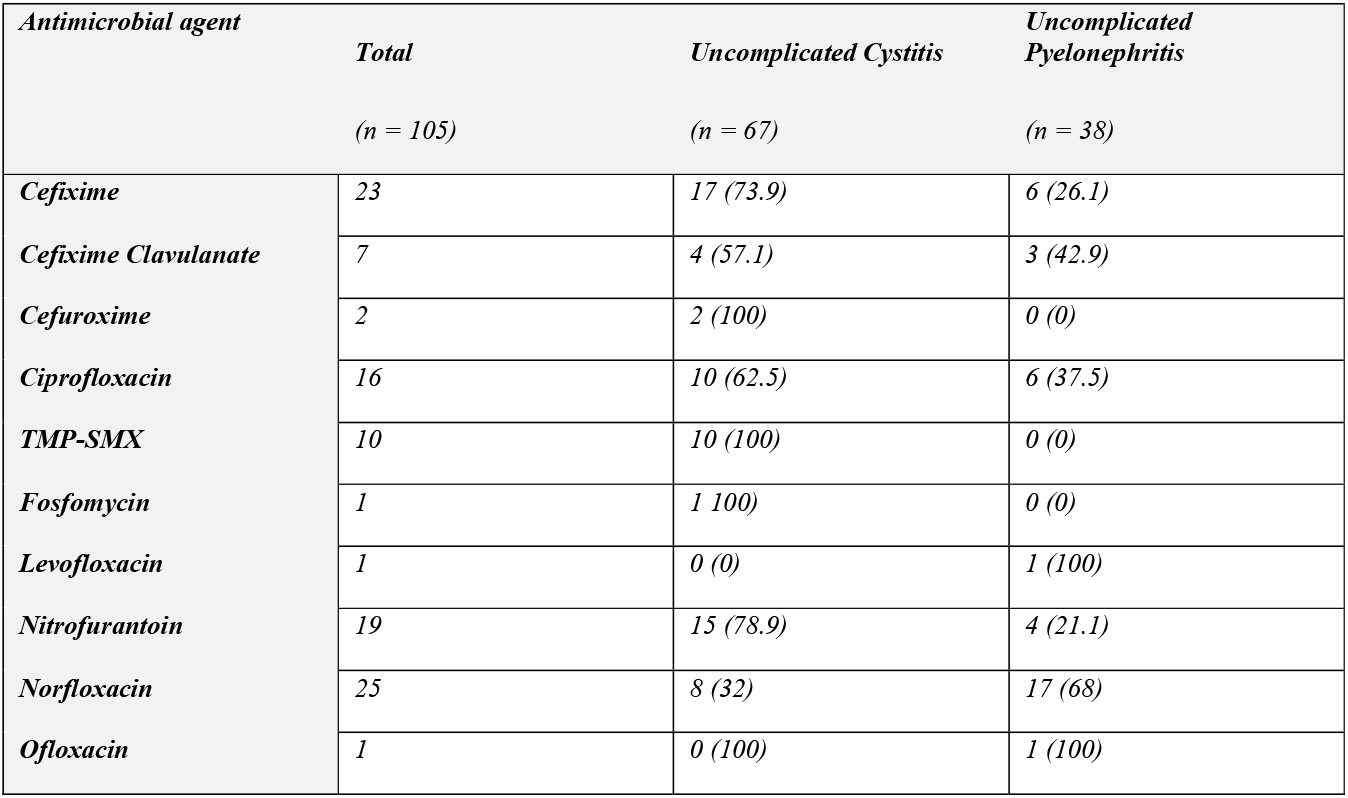
Antimicrobial agent prescribed in cases of Uncomplicated cystitis and Pyelonephritis.

| *Antimicrobial agent* | *Total*<br>*(n = 105)* | *Uncomplicated Cystitis*<br>*(n = 67)* | *Uncomplicated Pyelonephritis*<br>*(n = 38)* |
| --- | --- | --- | --- |
| *Cefixime* | 23 | 17 (73.9) | 6 (26.1) |
| *Cefixime Clavulanate* | 7 | 4 (57.1) | 3 (42.9) |
| *Cefuroxime* | 2 | 2 (100) | 0 (0) |
| *Ciprofloxacin* | 16 | 10 (62.5) | 6 (37.5) |
| *TMP-SMX* | 10 | 10 (100) | 0 (0) |
| *Fosfomycin* | 1 | 1 (100) | 0 (0) |
| *Levofloxacin* | 1 | 0 (0) | 1 (100) |
| *Nitrofurantoin* | 19 | 15 (78.9) | 4 (21.1) |
| *Norfloxacin* | 25 | 8 (32) | 17 (68) |
| *Ofloxacin* | 1 | 0 (100) | 1 (100) |

Parentheses contains percentages

*The duration of antimicrobial treatment has been summarized in Table 3. And the choice of antimicrobial agent was appropriate in 60 (57%) cases and duration in 30 (28*.*5%) cases*.

**Table 3.** Duration of antimicrobial agent in cases of Uncomplicated cystitis and Pyelonephritis. Represented in mean ± SD

| **Antimicrobial therapy** | **Total**<br>**(in days)** | **Uncomplicated Cystitis**<br>**(in days)** | **Uncomplicated Pyelonephritis**<br>**(in days)** |
| --- | --- | --- | --- |
| **Beta Lactams** | 5.1 ± 0.6 | 5.1 ± 0.7 | 5 |
| **Fluoroquinolones** | 5 ± 0.9 | 4.9 ± 0.7 | 5.1 ± 1 |
| **TMP-SMX** | 5.4 ± 0.8 | 5.4 ± 0.8 | 0 |
| **Fosfomycin** | 1 | 1 | 0 |
| **Nitrofurantoin** | 7.4 ± 1.2 | 7.3 ± 1.8 | 7.8 ± 1.3 |

The antimicrobial resistance rates are shown in Figure 3.

**Figure 3.**
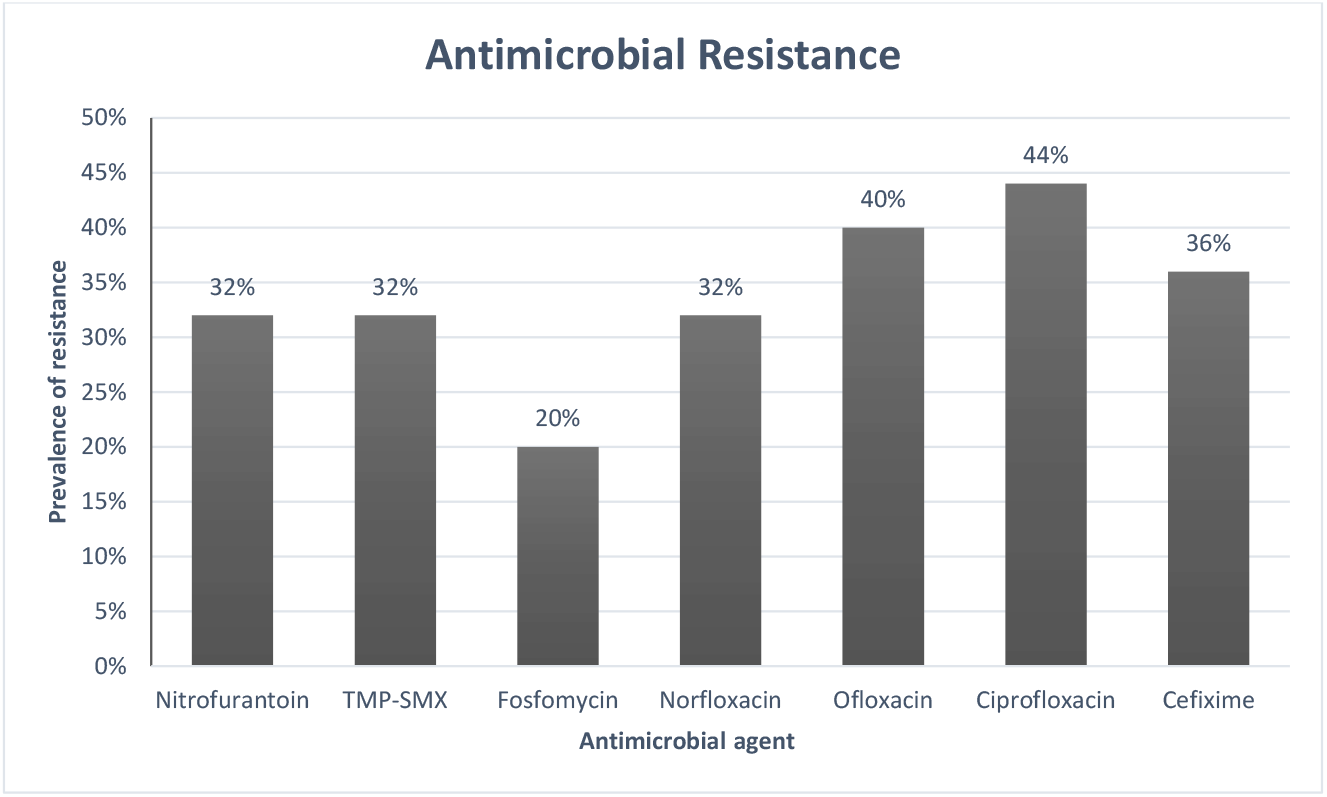
Antimicrobial resistance. Susceptibility Data showing the prevalence of antimicrobial resistance in our study

Upon follow-up, 92 participants (89.3%) had attained clinical cure out of which majority were cases of uncomplicated cystitis (67.4%). 11 (10.7%) participants had required escalation of treatment out of which, most were cases of uncomplicated pyelonephritis (72.7%). The clinical outcomes in relation to antimicrobial therapy is depicted in Table 4.

**Table 4.** Clinical outcomes in relation to type of UTI. Parentheses contains percentages

| Clinical Outcomes | Total<br>(n = 103) | Uncomplicated Cystitis<br>(n = 65) | Uncomplicated Pyelonephritis<br>(n = 38) | Chi- square test<br>(p < 0.05) |
| --- | --- | --- | --- | --- |
| Clinical Cure | 92 | 62 (95.4) | 30 (78.9) | 0.009 |
| Escalation of Treatment | 11 | 3 (4.6) | 8 (21.1) |  |

## Discussion

This study highlights current antimicrobial prescribing patterns in our hospital and the appropriateness of therapy as per IDSA guidelines with relation to the clinical outcomes. Appropriate use of antimicrobials is pivotal in reducing its ecological adverse effects and this is guided by local antibiotic susceptibility data.

Most of the patients (41%) in our study had received Fluoroquinolones as empirical therapy, even in cases of uncomplicated cystitis with duration of treatment being inappropriate. Only 38.8% of cases of uncomplicated cystitis had received the appropriate antimicrobial agent and 38.8% for the appropriate duration.

In a study done in the United States over a 9-year period the investigators demonstrated a similar trend.[9] Investigators studying antimicrobial prescribing patterns in North India in 2019 revealed that the most prescribed antimicrobials were quinolones and Beta-Lactam agents, and for extended duration.[10]

Multiple studies done in our setting demonstrate increasing antibiotic resistance against commonly prescribed antimicrobials such as fluoroquinolones, and beta lactams.[11–14] In our study, Fluoroquinolones and Beta lactams were prescribed more commonly in cases of uncomplicated cystitis which was not in accordance with IDSA guidelines. Similar trends in antimicrobial use in the outpatient setting have been demonstrated in studies based in the US.[15,16]

The spectrum of uropathogens and their antibiotic susceptibility data have important applications in guiding empirical therapy as per local antibiograms. In our study, the most commonly isolated uropathogen was E. coli, and the prevalence of MDR organisms was 36% which is in concordance with other studies showing a similar picture. This rise in MDR organisms has been attributed to the indiscriminate use of broad-spectrum antibiotics, especially for community acquired infections.

A similar study done in North India revealed the most common isolate was E. coli (61%). The prevalence of ESBL producing strains was 42% out of which majority was E. coli (34.4%)[14].

The antibiotic resistance data revealed greater levels of resistance among fluoroquinolones and Beta lactam group of antibiotics compared to NFT, TMP-SMX and Fosfomycin. These drugs have been studied and found to have excellent in-vitro activity against common uropathogens, especially E. coli and found to have high clinical cure rates.[17–19] However, the increased use of Fluoroquinolones in the outpatient setting have contributed to greater antibiotic resistance.

In a similar study done in South India, the most common uropathogen isolated was E. coli (70%). The antibiotic susceptibility data revealed that NFT was effective against both gram negative and gram positive uropathogens. However, Ciprofloxacin had shown overall decreased susceptibility.[15]

Other investigators observed that the most common organism isolated in their study was E. coli (36.11%) with high susceptibility to NFT (72.3%).[12]

Despite levels of TMP-SMX resistance to be greater than 20% in our study, it demonstrated a high clinical cure rate.

In our study, the frequency of clinical resolution in patients who were treated with NFT, TMP-SMX or Fosfomycin (96.7%) and Fluoroquinolones and Beta lactams (84.2%) were comparable.

A retrospective study done in a Spanish hospital over 7 years also revealed similar findings with high susceptibility rates to NFT, TMP-SMX and Fosfomycin.[20] which it also recommended as first choice for the treatment of uncomplicated cystitis.

The strengths of our study were that it was focused exclusively on persons visiting the outpatient department, which contrasts with other studies which also included in-patients. This avoids the inclusion of cases of complicated and recurrent UTI. The antibiotic susceptibility data obtained is not skewed by inclusion of specimens of in-patient cases which would portray a greater prevalence of Multidrug resistant organisms. Also, the case definitions and inclusion criteria are in accordance with international guidelines and the appropriateness of antimicrobial regimen by class and duration has not been studied in any other study in this setting.

There is an urgent need to procure more antibiotic susceptibility data from the community to guide empirical antimicrobial selection. This would reduce the development of antimicrobial resistance, incidence of recurrent UTI and hospitalization rates.

With rising antimicrobial resistance rates, adherence to the revised IDSA 2010 guidelines in the South Indian setting would improve antibiotic stewardship without compromising on clinical resolution in cases of Uncomplicated UTI.

## Conclusion

The current antimicrobial prescribing practices are grossly not in accordance with IDSA 2010 guidelines in terms of agent or duration in cases of Uncomplicated UTI, especially in case of Uncomplicated cystitis.

Use of NFT, TMP-SMX and Fosfomycin must be encouraged in the presence of favourable antibiotic susceptibility rates and non-inferior clinical cure rates.

Attempts at generating local susceptibility data must be encouraged in order to guide empirical therapy and monitor the rise of multi drug resistant organisms.

## Data Availability

All data produced in the present work are contained in the manuscript

## References

1. Schappert SM, Rechtsteiner EA. Ambulatory medical care utilization estimates for 2007. Vital Health Stat 13. 2011 Apr;(169):1–38.

2. Foxman B, Brown P. Epidemiology of urinary tract infections: Transmission and risk factors, incidence, and costs. Infectious Disease Clinics of North America. 2003;17(2):227–41.

3. Foxman B, Gillespie B, Koopman J, Zhang L, Palin K, Tallman P, et al. Risk factors for second urinary tract infection among college women. American Journal of Epidemiology. 2000;151(12):1194–205.

4. Kodner C, Gupton E. Recurrent Urinary Tract Infections in Women: Diagnosis and Management. American Family Physician. 2010;82(6):638–43.

5. Epp A, Larochelle A, Lovatsis D, Walter JE, Easton W, Farrell SA, et al. Recurrent Urinary Tract Infection. Journal of Obstetrics and Gynaecology Canada. 2010;32(11):1082–90.

6. Hooton TM. Recurrent urinary tract infection in women. International Journal of Antimicrobial Agents. 2001;17(4):259–68.

7. Hooton TM. Uncomplicated Urinary Tract Infection. New England Journal of Medicine. 2012 Mar 15;366(11):1028–37.

8. Gupta K, Hooton TM, Naber KG, Wullt B, Colgan R, Miller LG, et al. International clinical practice guidelines for the treatment of acute uncomplicated cystitis and pyelonephritis in women: A 2010 update by the Infectious Diseases Society of America and the European Society for Microbiology and Infectious Diseases. Clinical Infectious Diseases. 2011;52(5):103– 20.

9. Kobayashi M, Shapiro DJ, Hersh AL, Sanchez G v, Hicks LA. Outpatient Antibiotic Prescribing Practices for Uncomplicated Urinary Tract Infection in Women in the United States, 2002-2011. Open forum infect dis [Internet]. 2016 Aug 2 [cited 2020 Nov 14];3(3):ofw159–ofw159. Available from: https://pubmed.ncbi.nlm.nih.gov/27704014

10. Lakhani J, Lakhani S, Meera S, Sanket P, Sandeep J. Appropriate use of antimicrobial agents in urinary tract infections: Perception of physicians and resident doctors. Journal of Integrated Health Sciences. 2019;7(1):19–24.

11. Kalal BS, Nagaraj S. Urinary tract infections: A retrospective, descriptive study of causative organisms and antimicrobial pattern of samples received for culture, from a tertiary care setting. GERMS. 2016 Dec 2;6(4):132–8.

12. Patel H, Soni S, Bhagyalaxmi A, Patel N. Causative agents of urinary tract infections and their antimicrobial susceptibility patterns at a referral center in Western India: An audit to help clinicians prevent antibiotic misuse. Journal of Family Medicine and Primary Care. 2019 Jan;8(1):154–9.

13. Eshwarappa M, Dosegowda R, Aprameya IV, Khan MW, Kumar PS, Kempegowda P. Clinico-microbiological profile of urinary tract infection in South India. Indian Journal of Nephrology. 2011 Jan;21(1):30–6.

14. Akram M, Shahid M, Khan AU. Etiology and antibiotic resistance patterns of community-acquired urinary tract infections in J N M C Hospital Aligarh, India. Annals of Clinical Microbiology and Antimicrobials [Internet]. 2007 [cited 2020 Nov 14];6(1):4. Available from: https://doi.org/10.1186/1476-0711-6-4

15. George C, Norman G, Ramana Gv, Mukherjee D, Rao T. Treatment of uncomplicated symptomatic urinary tract infections: Resistance patterns and misuse of antibiotics. Journal of Family Medicine and Primary Care. 2015;4(3):416–21.

16. Khawcharoenporn T, Vasoo S, Singh K. Urinary Tract Infections due to Multidrug-Resistant Enterobacteriaceae: Prevalence and Risk Factors in a Chicago Emergency Department. Emergency medicine international [Internet]. 2013 Oct 31 [cited 2020 Nov 14];2013:258517. Available from: https://pubmed.ncbi.nlm.nih.gov/24307946

17. Sanchez G v., Baird AMG, Karlowsky JA, Master RN, Bordon JM. Nitrofurantoin retains antimicrobial activity against multidrug-resistant urinary escherichia coli from US outpatients. Journal of Antimicrobial Chemotherapy. 2014 Dec;69(12):3259–62.

18. Arredondo-García JL, Figueroa-Damián R, Rosas A, Jáuregui A, Corral M, Costa A, et al. Comparison of short-term treatment regimen of ciprofloxacin versus long-term treatment regimens of trimethoprim/sulfamethoxazole or norfloxacin for uncomplicated lower urinary tract infections: A randomized, multicentre, open-label, prospective study. Journal of Antimicrobial Chemotherapy. 2004 Oct;54(4):840–3.

19. Kavatha D, Giamarellou H, Alexiou Z, Vlachogiannis N, Pentea S, Gozadinos T, et al. Cefpodoxime-proxetil versus trimethoprim-sulfamethoxazole for short-term therapy of uncomplicated acute cystitis in women. Antimicrobial Agents and Chemotherapy. 2003 Mar;47(3):897–900.

20. Sorlozano A, Jimenez-Pacheco A, de Dios Luna Del Castillo J, Sampedro A, Martinez-Brocal A, Miranda-Casas C, et al. Evolution of the resistance to antibiotics of bacteria involved in urinary tract infections: A 7-year surveillance study. American Journal of Infection Control. 2014;42(10):1033–8.

